# Direct mailing of HPV self-sampling kits to older women non-participating in cervical screening in the Czech Republic

**DOI:** 10.1101/2023.10.05.23296387

**Authors:** Ondřej Ngo, Renata Chloupková, David Cibula, Jiří Sláma, Lucie Mandelová, Karel Hejduk, Marián Hajdúch, Petr Minka, Vladimíra Koudeláková, Hana Jaworek, Markéta Trnková, Peter Vaněk, Vladimír Dvořák, Ladislav Dušek, Ondřej Májek

**Author notes:** Corresponding author Ondřej Májek, Institute of Biostatistics and Analyses, Faculty of Medicine, Masaryk University, Kamenice 126/3, Brno, 625 00, Czech Republic.

## Abstract

**Background:** A population-based cervical cancer screening programme is implemented in the Czech Republic. However, participation is insufficient among women over 50 years. This study aimed to estimate the potential improvement in participation through directly mailed HPV self-sampling kits (HPVssk) compared to standard invitation letters in women aged 50-65 non-participating in screening.

**Methods:** The study recruited 1,564 eligible women (no cervical cancer screening in the last 3 years or more, no previous treatment associated with cervical lesions or cervical cancer). Eight hundred women were mailed with an HPVssk (HPVssk group), and 764 women were sent a standard invitation letter (control group) inviting them to a routine screening (Pap test). The primary outcome was a comparison of the overall participation rate between study groups using a binominal regression model.

**Results:** The participation rate in the HPVssk group was 13.4% (95% CI 11.2–15.9%; 7.4% of women returned the HPVssk and 6.0% attended gynaecological examination) and 5.0% (95% CI 3.6–6.8%) in the control group. Using the binominal regression model, the difference between the groups was estimated as 7.6% (95% CI 5.0-10.2%; p < 0.001). In the HPVssk group, 22% of women who returned HPVssk had a positive result and 70% of them underwent a follow-up examination.

**Conclusions:** Compared to traditional invitation letters, the direct mailing of the HPVssk achieved a significantly higher participation rate, along with a notable HPV positivity rate among HPVssk responders. This approach offers a potentially viable method for engaging women who have not yet attended a cervical screening programme.

## INTRODUCTION

Cervical cancer is a preventable disease thanks to the availability of HPV vaccination and screening; however, it is still the fourth most commonly diagnosed cancer and the fourth most common cancer cause of death in women worldwide, with more than 600 thousand new cases and 340 thousand deaths estimated for 2020 (1). Cervical cancer is a major health problem especially in less-resourced countries; however, it is also prevalent in hard-to-reach populations in high-income countries (2–4). For these reasons, the World Health Organization has initiated a global strategy to eliminate cervical cancer (5).

Cervical cancer screening is essential to reduce the population burden of cervical cancer. However, to be effective, it is essential to reach the maximum target population of women, especially those who do not regularly participate in screening. As the overwhelming cause of cervical cancer is infection with high-risk human papillomavirus (hrHPV), HPV self-sampling kits represent the most appropriate methodology to reach an underscreened population (4,6). Self-sampling tests represent a validated alternative to clinically collected specimens for HPV testing (7–9) and are a widely accepted and feasible method of testing (10,11).

HPV self-sampling kits have already been introduced as a primary screening method or are offered to non-participating women. They can increase participation in cervical screening by reducing barriers associated with clinical examination (12–15). Some studies have suggested that HPV self-sampling testing may be cost-effective if it increases screening attendance. Cost-effectiveness is also improved by reducing the cost of HPV self-testing, higher test sensitivity, and attracting never-tested and long-term undertested women (16,17).

Since 1960, pap smear testing has been performed in the Czech Republic as part of an annual preventive check-up where a sample is taken from the uterine cervix by a primary care gynaecologist. In 2008, the programme became an organised nationwide screening programme (all adult women to have regular Pap smear tests at 1-year intervals). At the same time, a network of cervical screening laboratories was established (18,19). In the Czech screening programme, the HPV test is also performed to classify the oncological risk in patients with mild and unclear cytological abnormalities and as a test to demonstrate the success of surgical treatment of cervical lesions. Since 2021, HPV co-test has been recommended and reimbursed as a part of the screening examination together with a cytological examination in all women aged 35 and 45 years (20).

Since 2014, a personalised invitation to cancer screening programmes for non-attenders has been introduced in the Czech Republic. All eligible individuals (up to the age of 70), who had not regularly attended the screening, have been invited by letter from the health insurance company; in the case of cervical screening, women are advised to visit their gynaecologist. If they do not respond to the invitation, they are invited again by letter after one year (21).

The annual coverage by examinations of cervical cancer screening reaches almost 60% in the Czech Republic. However, in women aged 50 years and older, screening coverage decreases with advancing age and, at the same time, the highest incidence of advanced cervical cancer is observed in older women (22,23). The initial data from pilot studies conducted in the Czech Republic, which utilized self-sampling kits for HPV detection across diverse target populations, revealed high levels of satisfaction among the individuals tested, as well as a successful rate of HPV detection (24–26).

Our study aimed to investigate the potential for increasing the overall participation rate of the target population of elderly women through cervical cancer screening using centralised direct mailing of self-sampling kits for the detection of high-risk HPV compared to standard invitation letters.

## METHODS

### Study design

The study was carried out in cooperation with a health insurance company (RBP, health insurance company) and used an already established system and algorithm for personalised invitation of women for cervical cancer screening in the Czech Republic. The health insurance company invites all insured women fulfilling the eligibility criteria on the month of their birthday. The study was conducted in February and March 2021. All women eligible for personalised invitation were selected and allocated to one of the following groups in an approximate 1:1 ratio each month:

- direct mailing of the HPV self-sampling kit (HPVssk group)
- mailing of standard invitation letter (control group)

In the HPVssk group, the women were also informed about the possibility of participating in a screening programme through examination by a gynaecologist.

All study participants provided written informed consent. This study was performed in compliance with the Helsinki Declaration according to the study ethics proposal approved by the Ethics Committee of the Faculty of Medicine and Dentistry at Palacky University and the University Hospital in Olomouc (protocol no. 150/18).

### Study participants

The health insurance company selected all eligible women aged 50-65 years in February and March 2021 according to a personalised invitation algorithm implemented in the health insurance company’s information system in the Czech Republic. These women who had not participated in screening in the last three years and had not undergone therapeutic and curative medical procedures for cervicovaginal lesions or cervical cancer were eligible for participation. Women who had already been previously invited for screening without response were also approached.

A total of 1,564 women who met the entry criteria were selected. Women were invited in their birth month, and the number of eligible women was similar in both recruitment months. Each month, health insurance company identified eligible women from an information system (raw data without any sorting) and the first 400 women received an HPV self-sampling kit. A standard invitation letter was sent to the remaining women.

### HPV self-sampling testing process

The women in the HPVssk group were mailed the instructions for test collection, a study information leaflet and consent to participate in the study, along with an invitation letter (letter in the HPVssk group was different than in the control group) and the HPV self-sampling kit. The attached documents provided information on HPV, the risk of cervical cancer, the self-sampling kit and the benefits of this test (27). This allowed women to make an informed decision about their participation in the study. The kit contained a brush device (Evalyn Brush, Rovers Medical Devices) designed to self-collect a cervicovaginal sample to test for the presence of oncogenic types of human papillomavirus in women (28). The women sent the collected sample by prepaid return envelope to the laboratory (due to the larger size of the parcel, the women had to pick it up at the post office and after use send the letter at the post office).

At the laboratory, Evalyn Brush heads were suspended with 3 ml PreservCyt transport medium (Hologic, Inc.), DNA was isolated using Ribospin vRD (GeneAll, Korea) according to manufacturer protocol, the presence of high-risk HPV was detected and genotyped (14 genotypes of high-risk HPV) using Anyplex II HPV HR Detection Assay (Seegene Inc., Korea) according to manufacturer protocol. The results were sent directly to the women with a recommendation for further action (it was possible to choose the form of sending the mentioned information by e-mail or by post). The organisational scheme of the HPVssk group is described in more detail in Figure 1. A reminder letter was sent to women who did not send a self-sampling kit to the laboratory at the end of April 2021.

**Figure 1:**
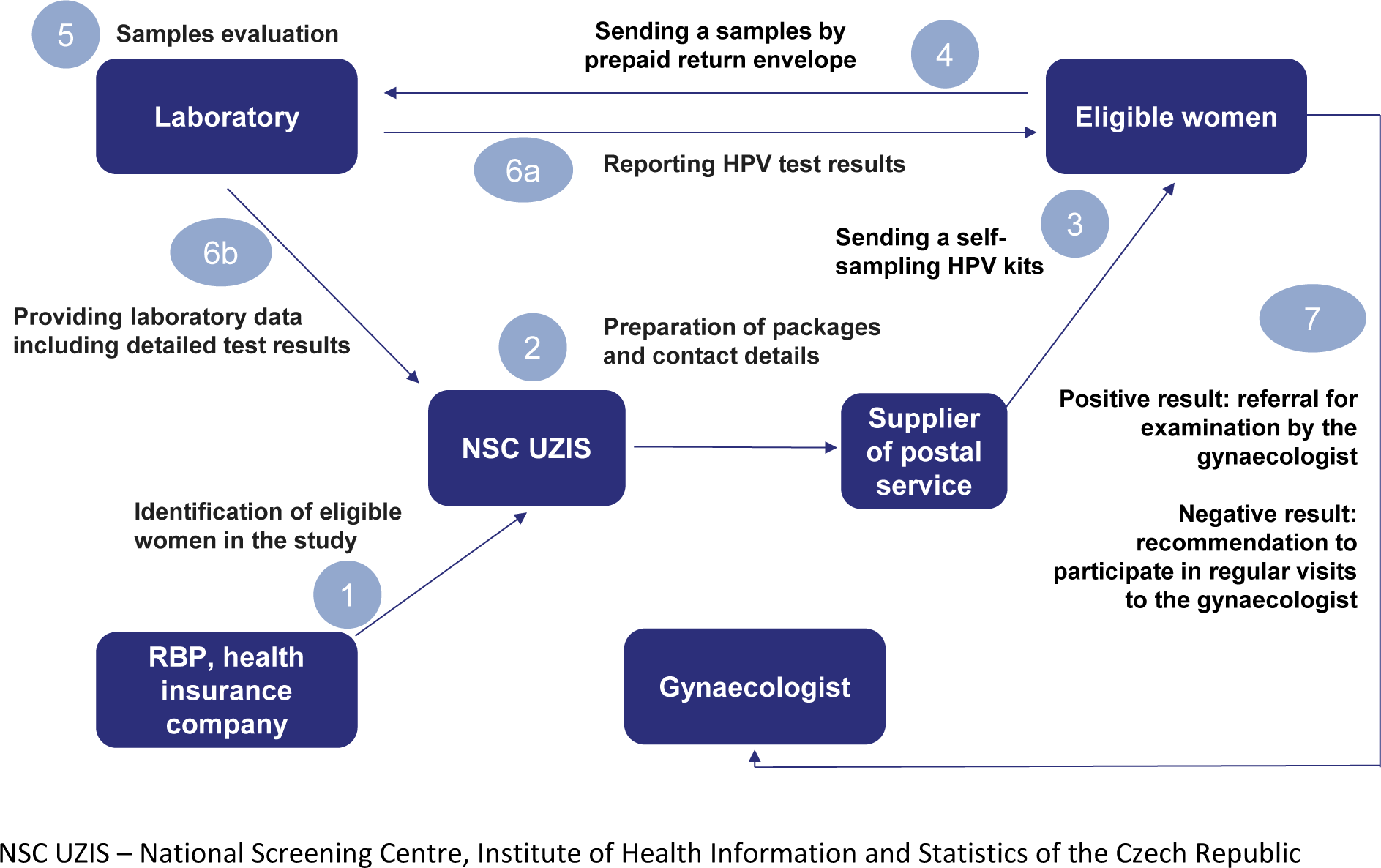
Scheme of the direct mailing of HPV self-sampling kits.

### Study Outcomes

The primary outcome of the study was to assess the overall participation rate after sending a personalised invitation to non-participating women and to compare the study groups in this endpoint. The overall participation rate for the HPVssk group considered either returning the HPV self-sampling kit to the laboratory or attending a screening examination by a gynaecologist. The overall participation rate is therefore defined as:

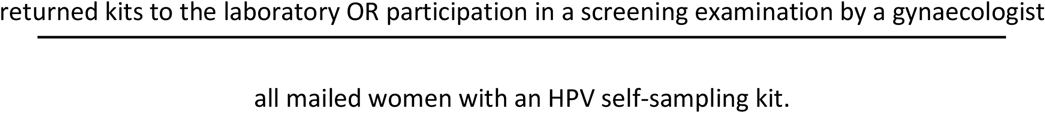

Attendance at the screening examination by a gynaecologist was monitored in the control group.

Secondary endpoints were comparisons of overall participation rates among different subgroups of women between study groups, the HPV positivity rate of women who sent an HPV self-sampling kit to the laboratory, and the proportion of follow-up examinations by a gynaecologist after a positive HPV test result.

### Sample size

Given the potential to reach approximately 800 women per month within the participating health insurance company, a power analysis was conducted to investigate if the power might be sufficient to compare overall participation rates between study groups. From the available results of the response to the standard invitation letters in the Czech Republic, we expected an overall participation rate for the control group of about 10%; an increase of 10 percentage points (29) due to the direct mailing of the self-sampling kits might have been achievable and significant from the public health viewpoint. Considering the implementation of the study in two months (approximately 1,600 invited women), a 5% significance level and the expected difference between the HPV and control group, we should achieve a statistical power of more than 90%.

### Data sources and statistical methods

Health insurance records for all women contacted in February and March 2021 were available and linked to the laboratory data (the results from the laboratory were managed separately in the electronic case-report forms ClinData of the Institute of Molecular and Translation Medicine and were made available for statistical analysis) for information on who sent back the HPV self-sampling kit. At the same time, the population-based registry (National Registry of Reimbursed Health Services, NRRHS) was used, which contained national data at the individual level on all reimbursed examinations. By linking to the NRRHS, it was possible to track women’s attendance at their preventive check-up by a gynaecologist in both groups and follow-up examinations after a positive result of the HPV self-sampling kit. In the HPVssk group, data were available on unclaimed and undeliverable letters from the postal service provider.

For the primary endpoint of the study, overall participation rates for both groups were analysed together for both batches. Samples sent to the laboratory and examinations performed by the gynaecologist within six months after the invitation were considered in the overall participation rate (numerator). All originally selected women were included in the denominator. This reflected the calculation of the overall participation rate in practice, where the exact number of undelivered letters is often unknown and the entire eligible population approached is reported as the denominator. Main statistics were supplemented with 95% confidence intervals (CI). Subsequently, the difference in overall participation rates between groups were assessed using a binomial regression model adjusted for number of invitations (represents how many times a woman has received an invitation; number 1 describes the first invitation to cervical cancer screening), letter variant (it describes screening programmes that the woman has not attended and was invited to by letter – cervical, breast and colorectal cancer screening) and age with the addition of a confidence interval for the difference and a p-value (a 5% significance level was considered). Pearson chi-square test was used to compare baseline characteristics (age, number of invitations and letter variant) between study groups and a 5% significance level was considered.

For the secondary endpoints, a subgroup analysis comparing study groups in overall participation rates (by age, number of invitations and letter variant) was performed using an adjusted binomial regression model. The proportion of women who tested positive for HPV (HPV 16, 18 and 45 were monitored separately) and the proportion of women who attended a gynaecological follow-up examination within six months after the positive result of the HPV self-sampling kit were calculated.

All statistical analysis was performed in the software tool Stata 15.

## RESULTS

### Participant flow and recruitment

A total of 1,564 eligible (assessed for eligibility according to the personalised invitation algorithm in the health insurance company information system) women were allocated into the two groups and all women were included in the final analysis. Eight hundred women were enrolled in the HPVssk group, 764 women in the control group. In the HPVssk group, 73 women did not pick up a letter at the post office. For 26 women, the letter was not deliverable for objective reasons (Figure 2).

**Figure 2:**
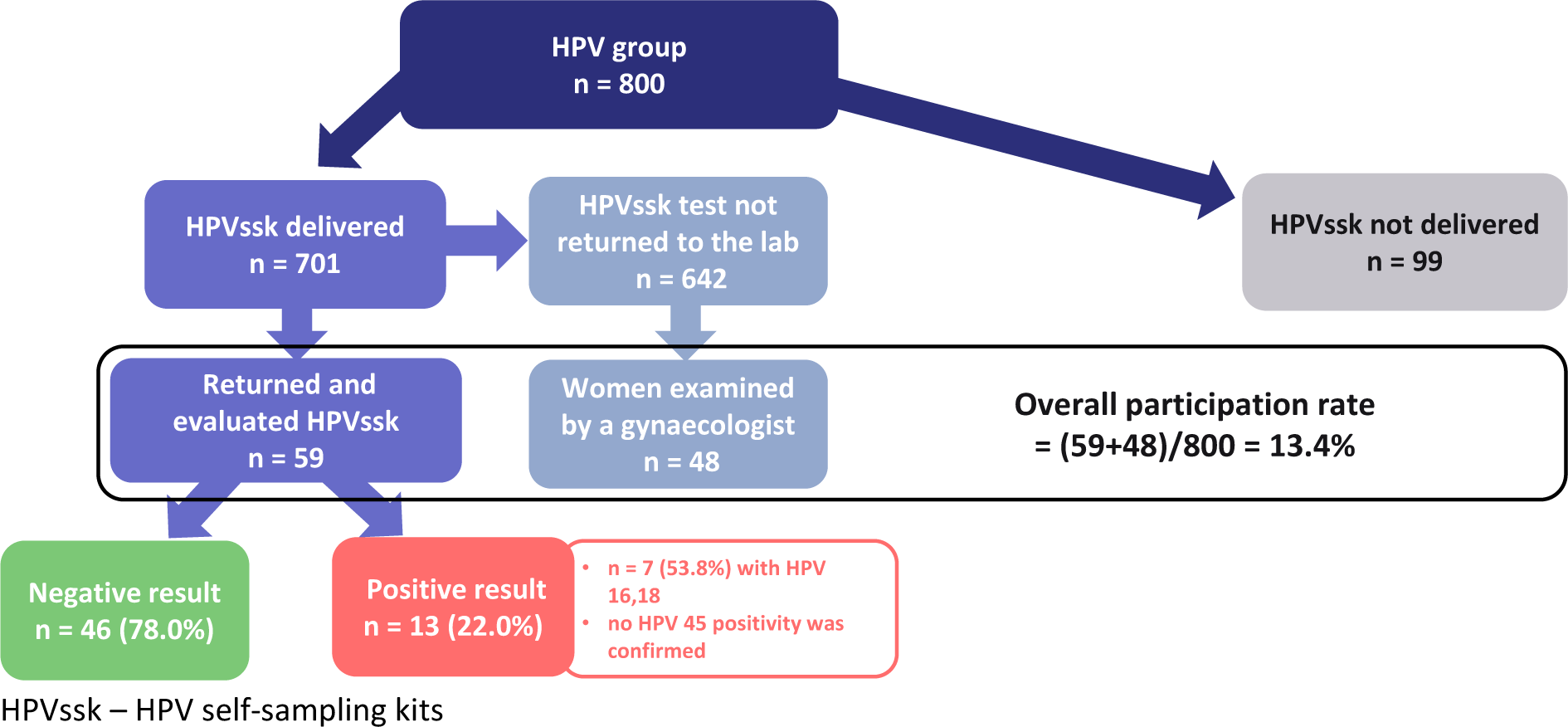
Flow diagram of women in the HPVssk group of the study.

### Baseline data

Within both groups, the highest representation of women was in the 60-65 age group, women who had not responded to two previous invitations, and women not participating in any cancer screening programme in the Czech Republic. There were no statistically significant differences between the study groups in the baseline characteristics (Table 1).

**Table 1:** Baseline characteristics of eligible population by study groups.

| Characteristics | HPVssk group<br>(HPV self-sampling test) |  | Control group<br>(standard invitation letter) |  | Comparison of the<br>study groups |
| --- | --- | --- | --- | --- | --- |
|  | No. | % | No. | % | p-value |
| Age group |  |  |  |  |  |
| 50–54 | 200 | 25.0% | 177 | 23.2% | 0.694 |
| 55–59 | 274 | 34.3% | 270 | 35.3% |  |
| 60–65 | 326 | 40.8% | 317 | 41.5% |  |
| No. of invitations<sup>a</sup> |  |  |  |  |  |
| 1 | 102 | 12.8% | 115 | 15.1% | 0.359 |
| 2 | 84 | 10.5% | 64 | 8.4% |  |
| 3 | 258 | 32.3% | 262 | 34.3% |  |
| 4 | 132 | 16.5% | 123 | 16.1% |  |
| 5 and more | 224 | 28.0% | 200 | 26.2% |  |
| Letter variant<sup>b</sup> |  |  |  |  |  |
| C | 94 | 11.8% | 100 | 13.1% | 0.231 |
| C and M | 180 | 22.5% | 190 | 24.9% |  |
| C and K | 73 | 9.1% | 81 | 10.6% |  |
| C and M and K | 453 | 56.6% | 393 | 51.4% |  |

a represents how many times a woman has received an invitation
b describes what screening programmes the invited women did not attend; C – cervical cancer screening, M – breast cancer screening, K – colorectal cancer screening
c comparison were made using the Pearson chi-square test

### Outcomes and estimation

The overall participation rate was 13.4% (95% confidence interval 11.2–15.9%) in the HPVssk group and 5.0% (95% confidence interval 3.6–6.8%) in the control group. In the HPVssk group, 59 (7.4%) women returned the self-sampling kit (all returned kits contained a sample and were valid for analysis in the laboratory) and another 48 (6.0%) directly attended the preventive gynaecological examination. The difference between the invitation methods was estimated to 7.6% (95% CI 5.0–10.2%; p < 0.001) in favour of the HPV self-sampling kits, as calculated using a binomial regression model adjusted for the number of invitations, letter variant, and age.

Overall participation rates among study groups by age, number of invitations, and letter variant were significantly higher in the HPVssk group in all subgroups compared to the control group (Table 2). The most considerable difference was observed among women who were invited for the first time or did not respond to the first invitation letter and were invited a second time. Another notable difference was observed for women in the study who were evaluated for invitation for cervical screening only or for cervical screening and one other cancer screening programme.

**Table 2:**
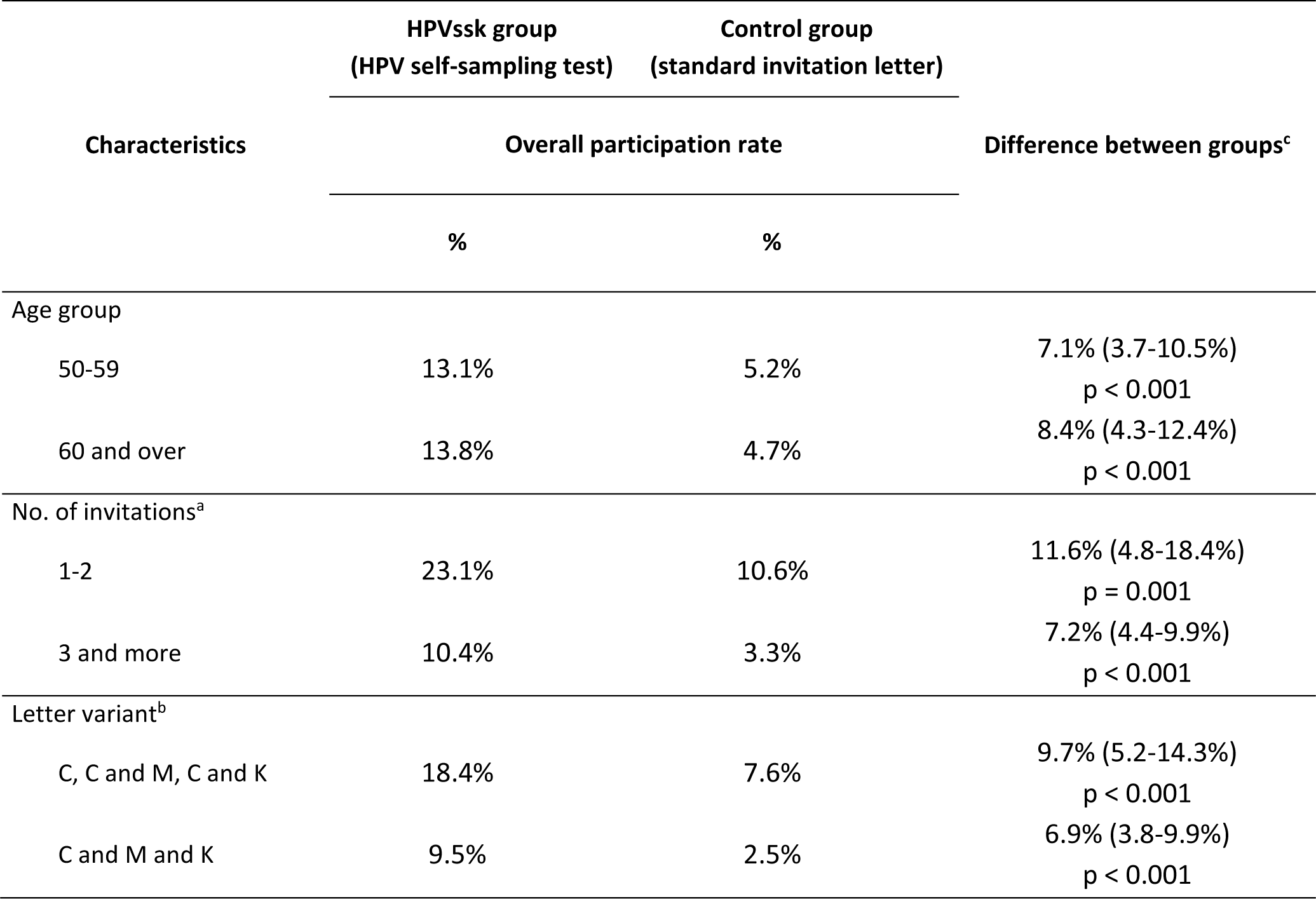
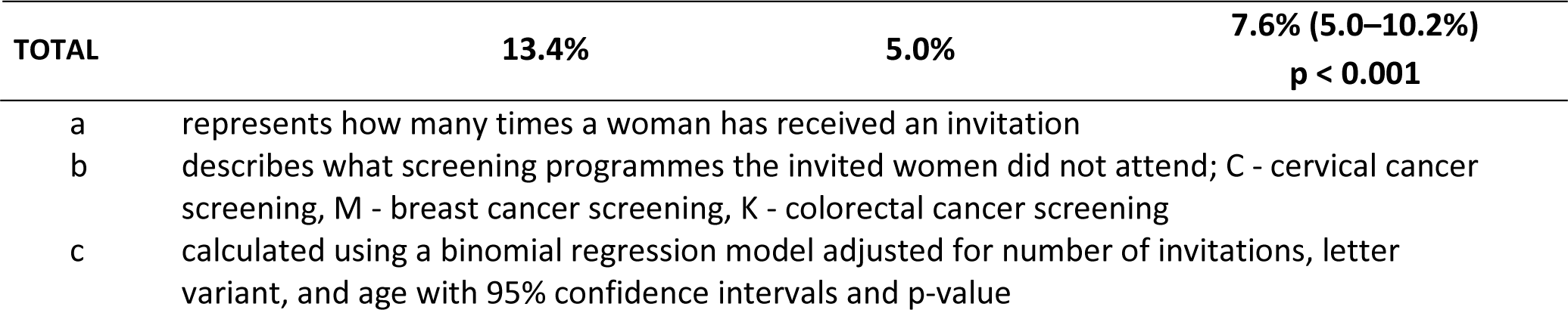
Comparison of overall participation rates between study groups by age, number of invitations and letter variant.

| Characteristics | HPVssk group<br>(HPV self-sampling test) | Control group<br>(standard invitation letter) | Difference between groups<sup>c</sup> |
| --- | --- | --- | --- |
|  | Overall participation rate |  |  |
|  | % | % |  |
| Age group |  |  |  |
| 50-59 | 13.1% | 5.2% | 7.1% (3.7-10.5%)<br>p < 0.001 |
| 60 and over | 13.8% | 4.7% | 8.4% (4.3-12.4%)<br>p < 0.001 |
| No. of invitations<sup>a</sup> |  |  |  |
| 1-2 | 23.1% | 10.6% | 11.6% (4.8-18.4%)<br>p = 0.001 |
| 3 and more | 10.4% | 3.3% | 7.2% (4.4-9.9%)<br>p < 0.001 |
| Letter variant<sup>b</sup> |  |  |  |
| C, C and M, C and K | 18.4% | 7.6% | 9.7% (5.2-14.3%)<br>p < 0.001 |
| C and M and K | 9.5% | 2.5% | 6.9% (3.8-9.9%)<br>p < 0.001 |
| **TOTAL** | **13.4%** | **5.0%** | **7.6% (5.0–10.2%)**<br>**p < 0.001** |

- a represents how many times a woman has received an invitation b describes what screening programmes the invited women did not attend; C - cervical cancer screening, M - breast cancer screening, K - colorectal cancer screening c calculated using a binomial regression model adjusted for number of invitations, letter variant, and age with 95% confidence intervals and p-value

The hrHPV examination was positive in 13 (22.0%) of 59 women returning the HPV self-sampling kits. Seven (53.8%) women were detected with HPV 16, 18 (no HPV 45 positivity was confirmed). All women with positive hrHPV were recommended to undergo a check-up with their gynaecologist. Nine women (69.2% of those who tested positive) underwent the follow-up examination (Figure 2).

## DISCUSSION

The results of our study suggest that the use of home-based HPV self-testing in a group of women who do not participate in screening can significantly improve their participation rates. The overall participation rate of women who were sent an HPV test was significantly higher than the participation rate of women who were approached by traditional invitation letter. The difference in participation rates was 7.6% (95% CI 5.0-10.2%; p < 0.001). In addition, higher overall participation rates were also observed for all subgroups of the HPVssk group compared with the control group, particularly for women who were invited for the first or second time, for cervical screening only, or for cervical screening plus one additional cancer screening program. The hrHPV test was positive in 22% (about half had HPV 16 and/or 18) of women in the HPVssk group who returned the self-sampling kit and approximately 70% of women arrived for a follow-up examination with a gynaecologist afterwards.

A number of meta-analyses have shown that offering HPV self-sampling kits can be a highly effective strategy for reaching never- or under-screened women compared to routine invitation and reminder letters to visit primary care provider (13,30,31). Incorporating this strategy into screening is therefore highly appropriate. At the same time, the initial results of a pilot study in the Czech Republic described a very good experience with HPV self-sampling kits (24) and our study has shown that direct mailing of self-selected HPV kits is feasible and acceptable to both patients and health care providers in the currently established system of personalised invitation in the Czech Republic.

In a meta-analysis published in 2018, the intention-to-treat (ITT) analysis (additionally including women approached with the HPV self-sampling kit who finally choose to have a clinical sample) showed a slightly higher difference in participation rate in favour of the HPV self-sampling kits than in our study. The pooled participation difference was 12.8% (95% confidence interval 10.4% to 15.1%) (4). More recent meta-analysis showed a pooled difference of 13.2% (95% CI 11.0%, 15.3%) in ITT analysis (13). In the aforementioned meta-analysis, the range of test positivity in studies with the HPV self-sample arm ranged from 5.7% and 29.4% with a pooled proportion of 11.1% (95% CI 10.0%, 12.2%) – our study observed a high positivity rate of 22%. Adherence of women to attend follow-up examination after a positive result of HPV self-sampling kits was observed and achieved similar results to our study – the pooled proportion from the meta-analysis was 79.0% (95% CI 67.9%, 88.3%).

The observed hrHPV positivity rate in our study in non-participating women aged 50-65 years was approximately three times higher compared to the group of women aged 35 and 45 years who were routinely screened by a gynaecologist for high-risk HPV co-test (positivity rate about 7%) in the Czech Republic (32). The study population is therefore at particular risk of developing cervical cancer, also due to the high prevalence of HPV 16 and HPV 18, where early detection of these HPV genotypes and regular follow-up can lead to prevention of cervical cancer development (33). It is known that HPV prevalence decreases with age (34) and it can be inferred, given the high positivity in our study, that the same method of testing in a resistant younger female population may lead to much higher hrHPV positivity.

Although the study did not comprise formal randomisation, the selection process lead to groups comparable in key variables. To further decrease confounding related to measured variables, all comparisons between group use adjustments with binomial regression modelling.

The women willing to return the kit for hrHPV testing needed to fill and sign the informed consent form and consent with personal data processing in the study. This may have been an important barrier to participation and we expect that paperless logistics within a regular screening offer may increase the participation rate. Another barrier to participation could have been picking up and returning the self-collection kit at the post office due to the COVID-19 pandemic (restriction of contact, more complicated logistics at post offices).

The study was performed in collaboration with one smaller health insurance company (the Czech health system includes 7 public health insurance companies in total). Nevertheless, we consider that the results are applicable for the entire Czech population. All health insurance companies have a uniform algorithm of personalised invitations in their information systems in the Czech Republic, so direct mailing of HPV self-sampling kits appears to be a feasible form of reaching non-participating women in this established invitation system.

The results of this study indicate a significantly positive impact of the offer of directly mailed self-sampling kits. Nevertheless, in addition to direct mailing in the Czech health care system, where screening programmes are based on regular check-ups with primary care physicians, it is also appropriate to consider other forms of the offering of self-sampling kits. There still remains a large proportion of women who have not responded even to direct mailing of HPV self-sampling kits, and therefore offering kits namely through health professionals may be another complementary way to increase participation among underscreened women (35,36).

In the context of the Czech Republic, with a long-standing organised cervical cancer screening programme and good geographical accessibility of primary care gynaecologists, the offer of directly mailed self-sampling kits for hrHPV examinations still leads to the significant increase of participation rate in elderly non-participating women. Women who are sent first invitations may respond even better to this form of invitation compared to standard invitation letter. HPV self-sampling kits represent a promising screening strategy to increase the participation of women who are under- or never-screened (13,37).

## Funding

This work was supported by the European Union within the framework of the European Social Fund, Operation Programme Employment [Optimisation of the cervical cancer screening programme by introducing HPV DNA detection by self-testing kits in women who do not attend the current screening programme in the long term, grant no. CZ.03.2.63/0.0/0.0/15_039/0008171]. We acknowledge the use of the research infrastructure for translational medicine (EATRIS-CZ, www.eatris.cz).

## Conflict of interest

The authors declare no conflict of interest. The funders had no role in the design of the study; in the collection, analyses, or interpretation of data; in the writing of the manuscript, or in the decision to publish the results.

## KeyPoints

- HPV self-sampling kit is a feasible and acceptable testing method for both patients and health care providers in an established system of personalised invitation.
- Direct mailing of HPV self-sampling kits to elderly non-participating women can lead to a significant increase in participation compared to invitation letters, especially among women who are newly invited to screening or participate in at least one other cancer screening programme.
- In the population of women over 50 years old who do not participate in cervical cancer screening, self-testing revealed a very high frequency of high-risk HPV positivity (including HPV 16/18 positivity), which may be associated with a similarly elevated incidence of cervical cancer.

## Data availability

The authors are not authorised to share any potentially identifiable patient level data. In justified cases, data may be formally requested through the corresponding author and the request for data will be assessed by the Institute of Health Information and Statistics of the Czech Republic.

## Notes

### Competing Interest Statement

The authors have declared no competing interest.

### Clinical Trial

NCT04226313

